# Autonomic dysfunction is associated with the development of arterial stiffness: The Whitehall II cohort

**DOI:** 10.1101/2022.02.24.22271384

**Authors:** Jonas Schaarup, Martin S. Christensen, Adam Hulman, Christian Stevns Hansen, Dorte Vistisen, Adam G. Tabák, Daniel R. Witte, Lasse Bjerg

**Author notes:** Corresponding author: Jonas Schaarup, Mail, Adresss: Steno Diabetes Center Aarhus, Hedeager 3, 2nd floor, 8200 Aarhus N, Denmark.

## Abstract

**Background and aim:** To examine the association between baseline level and change of autonomic nervous function with subsequent development of arterial stiffness.

**Methods:** Autonomic nervous function was assessed of 4,901 participants of the Whitehall II occupational cohort by heart rate variability (HRV) indices and resting heart rate (rHR) three times between 1997-2009, while arterial stiffness was assessed by carotid-femoral pulse wave velocity (PWV) measured twice between 2007-2013. First, individual HRV/rHR levels and annual changes were estimated. Then, we modelled the development of PWV by HRV/rHR using linear mixed effect models. First, we adjusted for sex and ethnicity (model 1), and then for socioeconomic and lifestyle factors, various clinical measurements, and medications (model 2).

**Results:** A decrease in HRV and unchanged rHR was associated with subsequent higher levels of PWV, but the effect of a change in HRV was less pronounced at higher ages. A typical individual aged 65 years with a SDNN level of 30 ms and a 2% annual decrease in SDNN had 1.32 (0.95; 1.69) higher PWV compared to one with the same age and SDNN level but with a 1% annual decrease in SDNN. Further adjustment had no major effect on the results.

**Conclusion:** People who experience a steeper decline of autonomic nervous function have higher levels of arterial stiffness. However, the association was weaker at higher ages.

## 1 Introduction

According to the World Health Organization, cardiovascular disease (CVD) is the leading cause of morbidity and mortality worldwide (1). CVD mortality markedly declined in high-income countries through the adoption of preventive measures and more targeted and effective risk factor management and treatment (2). Intermediate end-points on the causal pathway towards CVD are of high interest as they may identify individuals with elevated CVD risk at an earlier time-point than the conventional CVD risk stratification tools (3) and may open the opportunity for early management and treatment in those who have highest CVD risk.

Arterial stiffness is increasingly acknowledged as a subclinical indicator of cumulative CVD risk and also as an intermediate end-point for CVD (4-7). The gold standard method for its evaluation is the assessment of aortic pulse wave velocity (PWV). PWV is determined non-invasively by measuring the speed at which the arterial pressure waves move through the descending aorta (8), a feature directly dependent on the stiffness of the aorta. Arterial stiffness is primarily determined by the composition of the structural components of the arterial wall (9). In youth, arteries are compliant and elastic (8). With aging, elastin fibres are gradually replaced with stiffer collagen fibres, leading to progressively less elastic arterial wall (10). This process can be accelerated by modifiable risk factors such as smoking, hypertension, dysglycaemia, and hyperlipidaemia (8). As such, arterial stiffness may act as a proxy of the summed exposure to cardiometabolic risk factors (11). However, arterial stiffness is also modulated dynamically under the control of direct local endothelial signals and studies have shown an association between sympathetic tone and increased arterial stiffness (12).

The autonomic nervous system plays an important role in adaptive changes of cardiac and vascular responsiveness to internal and external requirements (13). Abnormal function of the autonomic nervous system as seen in cardiac autonomic neuropathy (CAN) may lead to maladaptive vascular dynamics and an abnormally invariant heart rate. Heart rate variability (HRV) is a validated measure for assessing CAN (14). HRV is defined as the beat-to-beat variability heart rate that provides an estimate of the balance between the sympathetic and parasympathetic tone. HRV can be measured based on electrocardiographic traces (ECG)(13).

An association between autonomic dysfunction and increased arterial stiffness is well established in both type 1 diabetes and type 2 diabetes (15-18). Furthermore, studies have shown that autonomic imbalance is already present in individuals with prediabetes (19) and CAN is associated with lower insulin sensitivity in a individuals without diabetes (20). CAN is also a predictor of incident diabetes and vascular diseases (21). This suggests both that autonomic dysfunction not only exists in people with established diabetes, it may also be associated with arterial stiffness independently of hyperglycaemia.

Our hypothesis is that individuals with preserved autonomic nervous function have lower levels of arterial stiffness and slower progression of arterial stiffening. In this etiological longitudinal analysis, we will examine to which degree individual levels and change in autonomic nervous function are associated with the progression of arterial stiffness in a general population.

## 2 Materials and methods

### 2.1 Study population

The Whitehall II study is an occupational cohort that originally included 10308 (3413 women and 6895 men) British civil servants in the age range of 35-55 years at recruitment in 1985. The cohort was invited for a clinical examination approximately every 5 years and additionally received a questionnaire every 2-3 years. HRV and resting heart rate (rHR) were first measured in phase 5 (1997-1999) and further obtained in phases 7 (2002-2004) and 9 (22). PWV was first measured in phase 9 (2008-2009), considered as baseline in the current study, and then in phase 11 (2012-2013). The inclusion criteria in this investigation were having at least one measurement of HRV, rHR and PWV and complete information on covariates.

The Whitehall II study was reviewed and approved by the UK NHS Health Research Authority London-Harrow Ethics Committee and written informed consent was obtained from each participant at each examination phase. The study was conducted according to the principles of the Helsinki Declaration.

### 2.2 Exposure

Resting heart rate (rHR) was measured in 5-minute resting 12-lead ECG recordings obtained after 5 minutes of rest in the supine position. Then, the normal-to-normal (NN) sinus rhythm was determined from the recordings with an automated algorithm to identify R-R intervals without the presence of arrhythmias, ectopic beats and/or branch-blocks. These measurements were used to calculate HRV indices in the time and frequency domain (23). We included the HRV exposures of time domain: standard deviation of the NN interval (SDNN) and root mean square of successive differences (RMSSD), and frequency domain by using a Blackman-Tukey algorithm: low frequency (in the 0.04–0.15 Hz frequency band) (LF) and high frequency (in the 0.15–0.4 Hz frequency band) (HF). To account for cardiac automatism from concurrent heart rate, we included inter-beat interval corrected HRV (cHRV), an approach described in Van Roon et al. (24-26) (supplemental material). rHR was included as a control exposure to supplement the analysis.

### 2.3 Outcome

Aortic stiffness was characterised by aortic pulse wave velocity (PWV), which is calculated from the time between the ECG systole and the arrival of the pressure wave at the femoral and carotid measurement sites and the distance between these two measurement sites. Applanation tonometry is a validated method for assessing carotid-femoral PWV (SphygmoCor, Atcor Medical, Australia). PWV measurements were performed in a supine position after 10 minutes of rest. The aortic path length was determined with a tape measure by subtracting the carotid-sternal notch distance from the femoral-sternal notch distance. PWV was measured twice for each participant and the average was calculated (27). If the recordings differed by more than 0.5 m/s, a third measurement was performed and the average of the two closest measurements was used for the analysis.

### 2.4 Covariates

Self-administered questionnaires included information on categorical covariates such as smoking (never, former, current), socioeconomic status (administrative, professional/executive, clerical support), medication use (antihypertensive, cardiovascular, and antidiabetic medication), incidence of hypertension and other CVD), and continuous variables such as physical activity (hours of moderate to vigorous exercise) and alcohol use (units last week). Information on body mass index (BMI), waist-hip ratio, high-density lipoprotein (HDL), low-density lipoprotein (LDL), total cholesterol, triglycerides, haemoglobin A1c (HbA1c), oral glucose tolerance test (OGTT), and fasting plasma glucose (FPG) were collected as continuous covariates at clinical examination.

Diagnosis of diabetes was based on a combination of the 1999 and 2006 WHO guidelines (28, 29): FPG ≥7.0 mmol/l, 2-h postload plasma glucose (2-h PG) ≥11.1 mmol/l during an OGTT, HbA1c ≥6.5%, or self-reported diagnosis. The OGTT was not part of the study protocol at phase 11. Therefore, diagnosis at phase 11 is only based on FPG, HbA1c and self-report.

### 2.5 Statistical analysis

Descriptive analyses were performed by each study phase to characterize the distribution of continuous variables (median, 25^th^ & 75^th^ percentile) and frequencies (numbers, percentage) for categorical variables.

To examine individual-specific levels and changes in HRV/ rHR and their effect on PWV development, we used a 2-step analysis approach. *Step 1*: HRV and rHR trajectories were analysed separately using linear mixed-effect models to account for the repeated measurement structure in the data (30). The intercept (value for a given age) and slope (age) were included both as fixed and random effects. Person-specific levels and changes in HRV and rHR were estimated by combining these fixed and random effects. Levels were estimated for each participant’s age at phase 9 (study baseline). HRV values were log-transformed prior to analysis to obtain normally distributed model residuals. The person-specific HRV levels were transformed back to the original scale, while annual changes were expressed as a percentage. The estimated person-specific HRV/ rHR levels at the phase 9 baseline and changes prior to baseline were then used as the exposures in the second step. *Step 2*: A linear mixed effect model was used to analyse the association between person-specific HRV/rHR estimates (both level and change) and PWV trajectories at phase 9 and 11. To assess PWV age-trajectories, we included age in the model (as both fixed and random effect) and its interaction with the exposures (HRV or rHR level and change estimated in step 1). Two models were fitted with different degrees of adjustment: model 1 was adjusted for ethnicity and sex; model 2 was further adjusted for socioeconomic status (SES), BMI, smoking status, alcohol use, physical activity, levels of total cholesterol, triglycerides, HbA1c, systolic blood pressure, anti-hypertensive medication and glucose lowering medication. We present the development of PWV as 5-year trajectories based on typical combinations of age and corresponding HRV/ rHR levels at phase 9 and typical levels of annual change (Supplementary Table 1S and Fig. 1S).

As earlier studies suggest that HRV should be adjusted for concurrent rHR (31, 32), we conducted similar analysis using rHR corrected HRV indices: SDNN (cSDNN), RMSSD (cRMSSD), HF (cHF), LF (cLF). A subgroup analysis was performed including only those without diabetes. Hereby, the participants with diabetes (before phase 9 or 11) were excluded (this analysis was not adjusted for glucose lowering medication in model 2).

Complete case analyses were conducted. Analyses were performed using R version 3.6.2, using the *nlme* and *Epi* packages.

## 3 Results

From the entire cohort, 6412 (62%) participants had at least one measurement of HRV, 5069 (49%) participants among them also had at least one measurement of PWV, where 4901 (48%) had full information on covariates (**Figure 1**). Regarding HRV, 1071 (22%) had one measurement, 2312 (47%) had two, and 1518 (31%) had three. In total, 1494 (30%) had one PWV assessment and 3407 (70%) had two. In phase 5, the median (25^th^; 75^th^ percentile) age was 54.0 years (50.2; 59.6), 26% were women, and the median SDNN was 35.4 ms (26.6; 46.2). In phase 9, considered the baseline for our analyses, median PWV was 8.04 m/s (7.02; 9.44). The median interval for collection of data was 10.4 years (10.2; 10.7) for the exposures (phase 5 to 9) and 4.1 years (4.0; 4.2) for the outcomes (phase 9 to 11). Further characteristics of the participants are summarised by phase in **Table 1**. The subpopulation included 4207 participants, as 694 participants were diagnosed with diabetes before phase 9.

**Figure 1:**
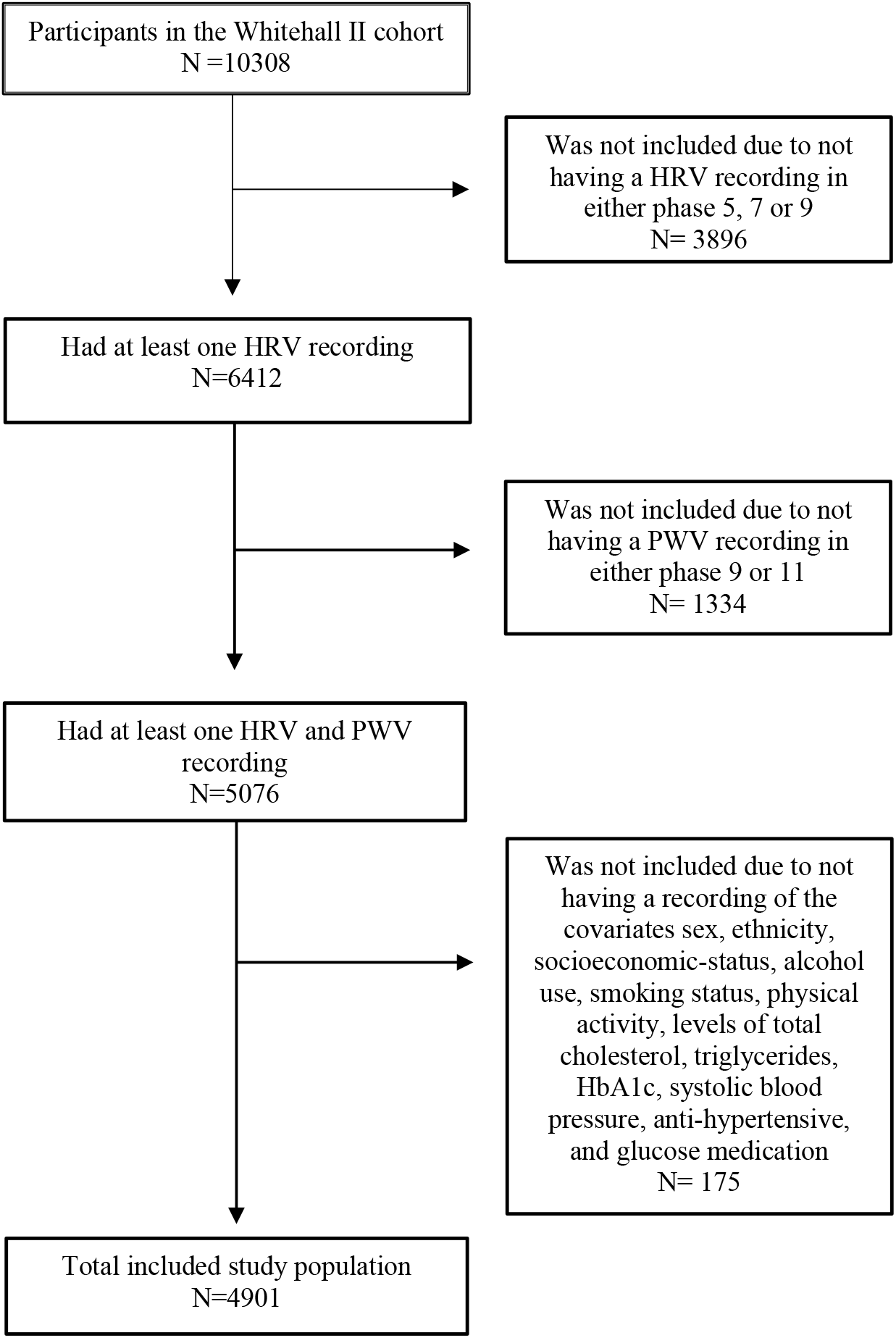
Flowchart of the included study population

**Table 1:** The study population characteristics by phase.

| **Table 1: The study population characteristics by phase** |  |  |  |  |
| --- | --- | --- | --- | --- |
|  | Phase 5 | Phase 7 | Phase 9 | Phase 11 |
| **Participation, N (%)** |  |  |  |  |
| Participated | 4715 (96.2) | 4681 (95.5) | 4870 (99.4) | 4649 (94.9) |
| Died before participation | 0 (0) | 0 (0) | 0 (0) | 92 (1.8) |
| Non-response /withdrawal | 186 (4.1) | 220 (4.5) | 31 (0.6) | 160 (3.3) |
| **Sex, N (%)** |  |  |  |  |
| Men | 3479 (73.8) | 3461 (73.9) | 3590 (73.7) | 3431 (73.8) |
| Women | 1236 (26.2) | 1220 (26.1) | 1280 (26.3) | 1218 (26.2) |
| **Ethnicity, N (%)** |  |  |  |  |
| White | 4364 (92.6) | 4334 (92.3) | 4500 (92.4) | 4317 (92.9) |
| Non-white | 351 (7.4) | 347 (7.4) | 470 (7.6) | 332 (7.1) |
| **Age (years)** | 54.0 (50.2; 59.6) | 59.5 (55.6; 65.1) | 64.4 (60.5; 70.0) | 68.5 (64.7; 74.1) |
| **BMI** | 25.4 (23.4; 27.7) | 25.9 (23.7; 28.2) | 25.9 (23.7; 28.5) | 25.9 (23.6; 28.6) |
| **Socioeconomic status N (%)** |  |  |  |  |
| Administrative | 1696 (36.0) | 1687 (36.0) | 1744 (35.8) | 1695 (36.5) |
| Professional/ Executive | 2418 (51.3) | 2398 (51.2) | 2498 (51.3) | 2380 (51.2) |
| Clerical/ Support | 601 (12.7) | 596 (12.7) | 628 (12.9) | 574 (12.3) |
| **Health behaviour determinants** |  |  |  |  |
| **Smoking N (%)** |  |  |  |  |
| Never | 2326 (51.3) | 2332 (50.2) | 2371 (48.9) | 2142 (47.5) |
| Former | 1844 (40.7) | 2001 (43.0) | 2229 (46.0) | 2224 (49.3) |
| Current | 364 (8.0) | 316 (6.8) | 248 (5.1) | 142 (3.1) |
| **Alcohol units per week** | 10 (4.0; 20) | 9 (3.0; 18.0) | 8 (2.0; 15.0) | 7 (2.0; 14.0) |
| **Physical Activity** hours/week moderate - vigorous physical activity | 12.4 (4.4; 23.1) | 13.5 (5.2; 24.7) | 12.8 (4.9; 24.9) | 12.8 (4.5; 24.7) |
| **Blood measurements** |  |  |  |  |
| HDL cholesterol (mmol/L) | 1.5 (1.2; 1.7) | 1.6 (1.3; 1.8) | 1.5 (1.3; 1.9) | 1.6 (1.3; 1.9) |
| LDL cholesterol (mmol/L) | 3.8 (3.2; 4.4) | 3.5 (2.9; 4.1) | 3.0 (2.3; 3.7) | 2.8 (2.2; 3.5) |
| Total cholesterol (mmol/L) | 5.8 (5.2; 6.5) | 5.7 (5.0; 6.4) | 5.2 (4.5; 5.9) | 5.1 (4.3; 5.8) |
| Triglycerides (mmol/L) | 1.1 (0.8; 1.4) | 1.1 (0.8; 1.6) | 1.0 (0.8; 1.5) | 1.0 (0.8; 1.4) |
| HbA1c (%) | - | 5.2 (5.0; 5.5) | 5.6 (5.4; 5.9) | 5.7 (5.5; 6.0) |
| Glucose: fasting (mmol/L) | 5.0 (4.7; 5.4) | 5.2 (4.9; 5.6) | 5.1 (4.8; 5.5) | 5.2 (4.9; 5.6) |
| Systolic blood pressure (mmHg) | 120.0 (110.3; 131.0) | 126.0 (115.0; 136.0) | 123.5 (113.5; 134.0) | 126.0 (116.0; 137.0) |
| Diastolic blood pressure (mmHg) | 77.0 (70.0; 84.0) | 73.0 (67.0; 80.0) | 70.5 (63.5; 77.0) | 70.5 (64.0; 77.0) |
| **Exposure** |  |  |  |  |
| rHR (resting bpm) | 67.7 (61.3; 75.1) | 66.4 (59.6; 73.6) | 66.2 (59.3; 73.6) | - |
| SDNN (ms) | 35.4 (26.6; 46.2) | 33.9 (25.8; 44.7) | 30.0 (22.2; 40.5) | - |
| cSDNN(cv) | 3.9 (3.1; 5.0) | 3.7 (2.5; 4.8) | 3.3 (2.5; 4.3) | - |
| RMSSD (ms) | 21.0 (14.1; 29.9) | 20.6 (14.0; 30.4) | 17.8 (12.0; 27.3) | - |
| cRMSSD (cv) | 2.33 (1.68; 3.23) | 2.23 (1.59; 3.16) | 1.92 (1.37; 2.86) | - |
| LF (ms<sup>2</sup>) | 334.1 (178.0; 608.111) | 286 (158.5; 527.8) | 224.4 (115.7; 448.8) | - |
| cLF (cv) | 4.24 (2.34; 7.18) | 3.49 (1.95; 6.15) | 2.70 (1.42; 5.19) | - |
| HF (ms<sup>2</sup>) | 224.4 (65.6; 265.6) | 117 (57.3; 242.6) | 88.2 (42.6; 191.1) | - |
| cHF (cv) | 1.69 (0.87; 3.18) | 1.37 (0.73; 2.71) | 1.06 (0.54; 2.15) | - |
| **Outcome** |  |  |  |  |
| PWV (m/s) | - | - | 8.04 (7.02; 9.44) | 8.53 (7.31; 10.27) |
| **Disease** |  |  |  |  |
| CVD, N (%) |  |  |  |  |
| Yes | 423 (8.7) | 409 (8.5) | 476 (9.5) | 404 (8.4) |
| No | 4449 (91.3) | 4449 (91.5) | 4547 (90.5) | 4402 (91.6) |
| Diabetes, N (%) |  |  |  |  |
| Yes | 272 (5.7) | 337 (7.2) | 483 (9.9) | 389 (8.4) |
| No | 4444 (94.3) | 4344 (92.8) | 4387 (90.1) | 4260 (91.6) |
| **Medication** |  |  |  |  |
| Anti-hypertensive medication, N (%) |  |  |  |  |
| Yes | 476 (10.2) | 950 (20.4) | 1598 (32.8) | 1880 (40.4) |
| No | 4206 (89.8) | 3712 (79.6) | 3268 (67.2) | 2768 (59.6) |
| CVD medication, N (%) |  |  |  |  |
| Yes | 602 (12.9) | 1202 (25.8) | 2391 (49.1) | 2693 (57.9) |
| No | 4080 (87.1) | 3460 (74.2) | 2475 (50.9) | 1955 (42.1) |
| Glucose lowering medication, N (%) |  |  |  |  |
| Yes | 51 (1.1) | 110 (2.4) | 188 (3.9) | 287 (6.2) |
| No | 4631 (98.9) | 4552 (97.6) | 4678 (96.1) | 4361 (93.8) |
| Characteristics describes the population who are participating in each phase<br>Categorical data shown in N (%)<br>Continuous data shown in median (25<sup>th</sup> percentile; 75<sup>th</sup> percentile) |  |  |  |  |

### 3.1 HRV / rHR levels and annual change

Model-based individual-specific HRV indices and rHR levels and annual change by age are summarised in **Table 1S** in the supplementary material. HRV decreased with age, and the HRV levels were lower in older age groups. E.g the annual median (25^th^ percentile; 75^th^ percentile) decrease for SDNN was -1.5% (−1.9; -1.1) irrespective of age and the median SDNN level for individuals aged below 65 years was 32.9 ms (28.5; 37.5), while for individuals aged above 70 years it was 27.4 ms (22.4; 33.6) (**Table 1S**). Furthermore, there was a strong correlation between estimated levels at phase 9 and annual change for both exposures, rHR and HRV (**Figure 1S**).

### 3.2 Association between level and change in HRV/ rHR estimates and the development of PWV

5-year PWV trajectories were estimated for a combination of typical HRV levels and changes based on the analysis in step 1 (**Figures 2 & 3**). We chose to show our results at the corresponding median HRV levels at age interval 60-65, 65-70 and 70-75 respectively and modelled two scenarios with regards to annual change (HRV with either a smaller or steeper decrease).

**Figur 2:**
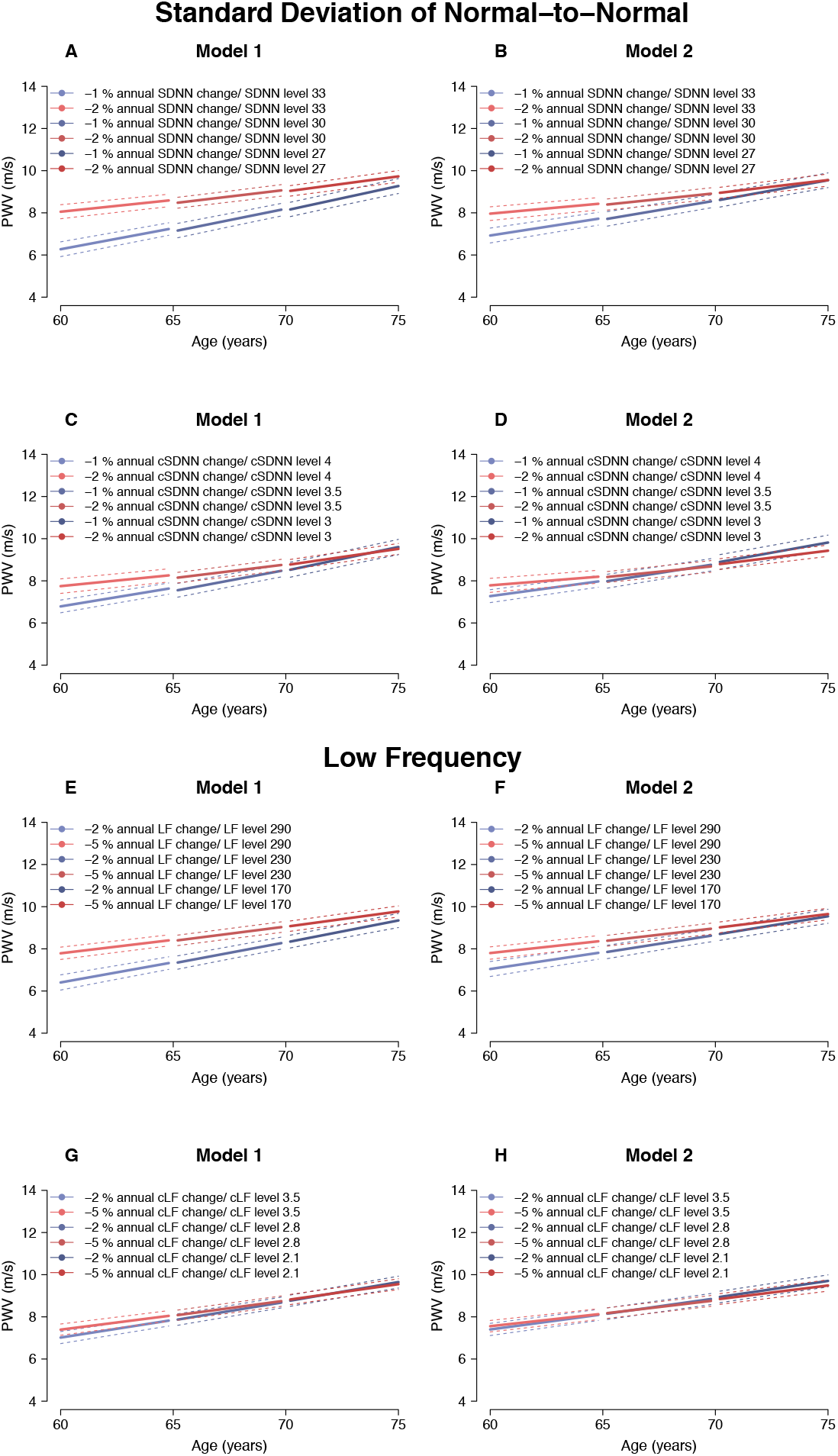
5-years PWV trajectories association with changes in HRV indices that mainly are characterized by mixed sympathetic and parasympathetic influences (SDNN and LF). The dotted lines show 95% confidence interval. A) Model 1: PWV trajectories for individuals with either -1% or -2% annual SDNN decrease B) Model 2: PWV trajectories for individuals with either -1% or -2% annual SDNN decrease C) Model 1: PWV trajectories for individuals with either -1% or -2% annual cSDNN decrease D) Model 2: PWV trajectories for individuals with either - 1% or -2% annual cSDNN decrease. E) Model 1: PWV trajectories for individuals with either -2% or -5% annual LF decrease F) Model 2: PWV trajectories for individuals with either -2% or -5% annual LF decrease G) Model 1: PWV trajectories for individuals with either -2% or -5% annual cLF decrease H) Model 2: PWV trajectories for individuals with either -2% or -5% annual cLF decrease. Model 1 (Sex= Men, Ethnicity= White), Model 2 (Sex=Male, Ethnicity=.White, SES= Professional/executive, BMI= 25, Smoking status= Non-smoker, Alcohol use= 8 units per week, Physical activity= 13 hours weekly of moderate to vigorous, Total cholesterol= 5.2mmol/L, Triglycerides= 1mmol/L, HbA1c= 5.6%, Systolic blood pressure= 124mmHg, Antihypertensive medication= Not using, Glucose lowering medication= Not using)

**Figure 3:**
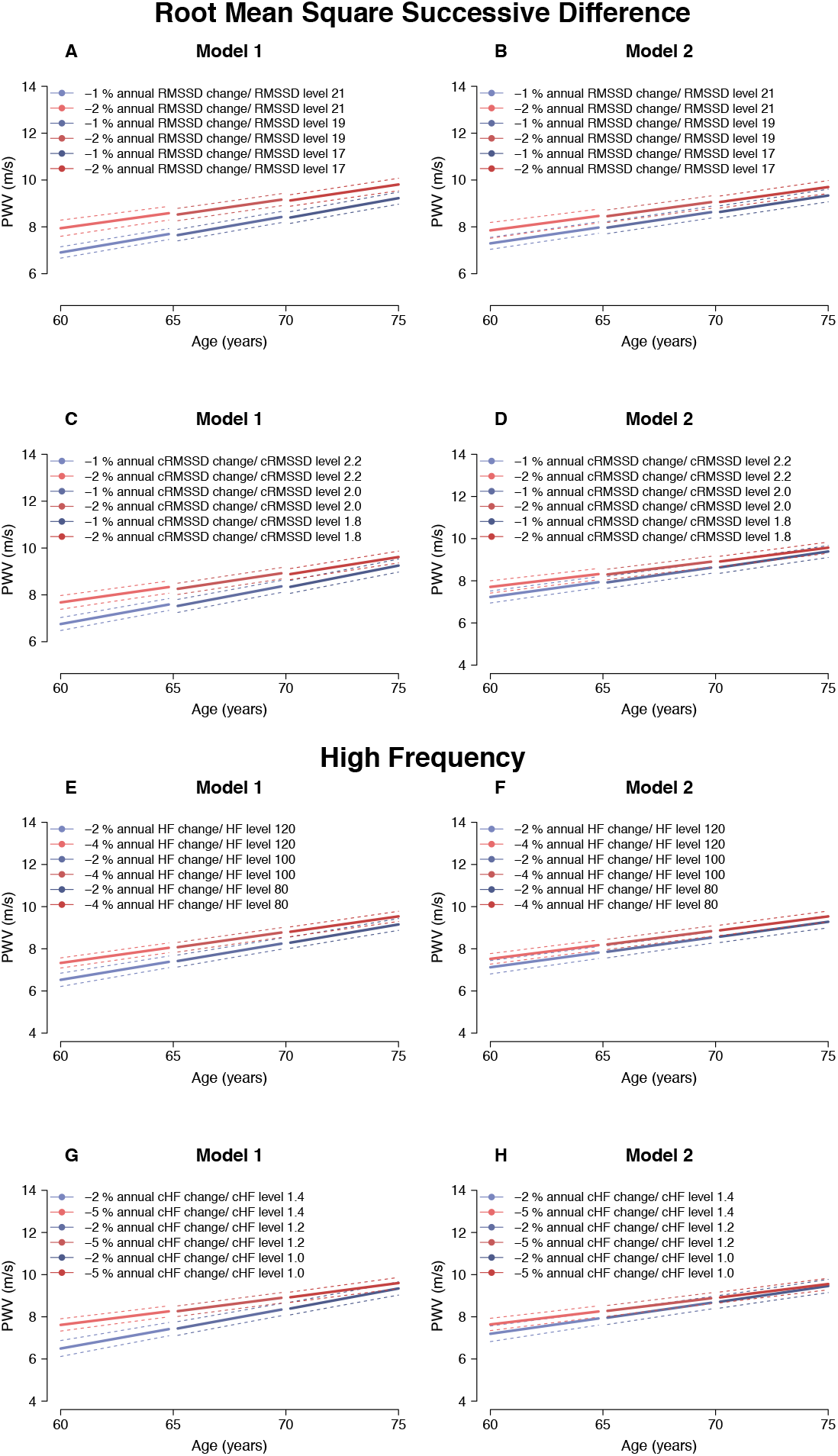
5-years PWV trajectories and its association with changes in HRV indices that mainly are characterized by parasympathetic influence (RMSSD and HF). The typical individuals have a baseline starting at 60, 65 or 70, respectively. The dotted lines show 95% confidence interval. A) Model 1: PWV trajectories for individuals with either - 1% or -2% annual RMSSD decrease B) Model 2: PWV trajectories for individuals with either -1% or -2% annual RMSSD decrease C) Model 1: PWV trajectories for individuals with either -1% or -2% annual cRMSSD decrease D) Model 2: PWV trajectories for individuals with either -1% or -2% annual cRMSSD decrease E) Model 1: PWV trajectories for individuals with either -2% or -4% annual HF decrease F) Model 2: PWV trajectories for individuals with either -2% or -4% annual HF decrease G) Model 1: PWV trajectories for individuals with either -2% or -5% annual cHF decrease H) Model 2: PWV trajectories for individuals with either -2% or -5% annual cHF decrease. Model 1 (Sex= Men, Ethnicity= White), Model 2 (Sex=Male, Ethnicity=. White, SES= Professional/executive, BMI= 25, Smoking status= Non-smoker, Alcohol use= 8 units per week, Physical activity= 13 hours weekly of moderate to vigorous, Total cholesterol= 5.2mmol/L, Triglycerides= 1mmol/L, HbA1c= 5.6%, Systolic blood pressure= 124mmHg, Antihypertensive medication= Not using, Glucose lowering medication= Not using)

**Figure 4:**
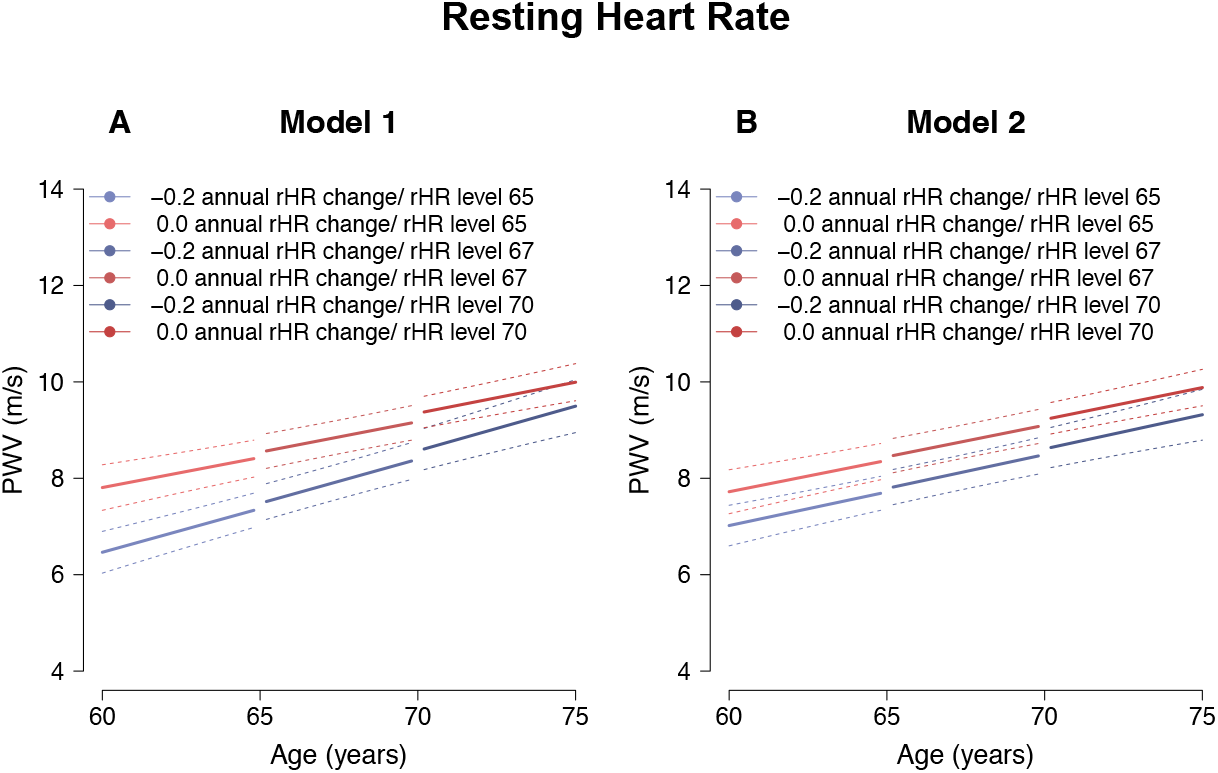
5-years PWV trajectories and its association with changes in rHR. The typical individuals have baseline age at 60, 65 and 70, respectively. The dotted lines show 95% confidence interval. The typical individuals have a baseline starting at 60, 65 or 70, respectively. The dotted lines show 95% confidence interval. A) Model 1: PWV trajectories for individuals with either 0% or -0.2% annual rHR change B) Model 2: PWV trajectories for individuals with either 0% or -0.2% annual rHR. Model 1 (Sex= Men, Ethnicity= White), Model 2 (Sex=Male, Ethnicity=. White, SES= Professional/executive, BMI= 25, Smoking status= Non-smoker, Alcohol use= 8 units per week, Physical activity= 13 hours weekly of moderate to vigorous, Total cholesterol= 5.2 mmol/L, Triglycerides= 1mmol/L, HbA1c= 5.6%, Systolic blood pressure= 124mmHg, Antihypertensive medication= Not using, Glucose lowering medication= Not using)

Those with a steeper decrease of HRV indices (SDNN, LF, RMSSD and HF) had a higher level of PWV than those with a less steep decrease (**Figure 2 & 3**). However, the effect of change in HRV indices was less pronounced at higher ages. E.g., based on model 1, the PWV difference between individuals with -2% and -1% annual change in SDNN was 1.78 m/s (95CI: 1.28; 2.28) at age 60 and 0.88 m/s (95CI: 0.55; 1.21) at age 70 (supplemental Fig. S3A and Table S2). Those with a steeper decrease in HRV had a slower annual increase in PWV than for those with a less pronounced decreasing HRV trajectory. Further adjustment (Model 2) had minor impact on these results (**Figure 2 & 3**). After the HRV indices were adjusted for their concurrent heart rate, the difference in PWV level was diminished (**Figure 2 & 3**).

In the sub-population without diabetes, the results were persistent in model 1, and after the further adjustments of model 2 (**supplemental Fig. S5c and Fig. S5d**). As this analysis, did not differ from the main population, it was not further investigated (Supplementary fig 5S.).

5-year PWV trajectories were estimated for a combination of typical rHR levels and changes based on the analysis in step 1 (Figure 3). We chose to show our results at rHR levels of 65, 67 and 70 bpm at ages 60, 65 and 70, respectively and modelled two scenarios with regards to change (−0.2 bpm/year and 0.0 bpm/year). PWV levels and changes are summarized in **Table 2**.

Those with no HR change had a higher level of PWV than those with -0.2 bpm/year rHR change regardless of age (**Figure 3 & Table 2**) The difference was 1.34 m/s (95CI: 0.56; 2.13) at age 60 and 0.77 m/s (95CI: 0.16; 1.37) at age 70 (**supplemental Fig. S4 and Table 2S**). rHR had a smaller effect on PWV slope than on level. Those with an increasing rHR (0.0 bpm/year) had a 0.12 m/s (95CI: 0.08; 0.16) annual increase in PWV, which was 0.06 (95CI: -0.02; 0.13) slower than for those with a decreasing rHR trajectory (−0.2 bpm/year). Further adjustment (Model 2) did not have a major effect on the results.

## 4 Discussion

In this study of 4,901 individuals, we showed how autonomic nervous function decreased with age, while resting heart rate tended to remains constant. Individuals with a steeper decline in autonomic nervous function or with an unchanged rHR had subsequent higher levels of arterial stiffness. However, this association was less pronounced at higher ages.

Several studies have found lower autonomic nervous function assessed by HRV indices to be associated with arterial stiffness in individuals with either type 1 or type 2 diabetes, suggesting that autonomic nervous function may play a mediating role in the association between diabetes and the development of arterial stiffness (15, 16, 18, 33, 34). Our finding of an association between autonomic nervous function and arterial stiffness in a general population and in a subgroup without diabetes, extends these findings and suggests a relation between autonomic nervous function and arterial stiffness also in the absence of diabetes.

In contrast to this study, previous studies have mainly been cross-sectional and did not examine the longitudinal association between level and change of autonomic nervous function with development of arterial stiffness (15, 16, 18, 33, 34). Two possible mechanisms might explain how the steeper decrease in autonomic nervous function is related to higher level of arterial stiffness. First, a decrease in autonomic nervous function may influence the elasticity of the arterial wall by increasing the vascular tone of large arteries. In rats, adequate autonomic nervous function is important in the maintenance of the elasticity in the aorta, suggesting that increased sympathetic activity can cause damage to elastin fibres, resulting in reduced elasticity (17, 34-36). However, results observed in animal models cannot be translated directly to human population studies.

Second, heart rate is under strong autonomic control, and autonomic dysfunction not only leads to lower heart rate variability, but also to a higher resting heart rate. A higher resting heart rate in turn increases arterial stiffness, likely through alterations in blood flow dynamics leading to higher shear stress (37). A previous study in a general population has shown that increased resting heart rate is associated with arterial stiffness (38), which is also supported by our findings of an association between rHR levels and its change, and the development of arterial stiffness. We are aware that there is an inverse relationship between rHR and HRV, indicating that HRV are not only affected by autonomic nervous function, but also cardiac automatism. We have attempted to accommodate the influence of concurrent rHR on HRV. After the adjustment of HRV indices that is influenced by both sympathetic and parasympathetic activity (SDNN and LF) for concurrent rHR the association between autonomic dysfunction and development of arterial stiffness was substantially diminished, whereas HRV indices influenced by parasympathetic activity (RMSSD and HF) was less affected. Still, there has not been developed a standardized method of adjusting HRV for cardiac automatism, (25) hence we present both rHR adjusted and non-adjusted HRV models.

We assessed potential covariates through literature to minimize the impact of confounding on the association between HRV and PWV considering sex, ethnicity, socioeconomic and lifestyle factors, various clinical and lab measurements as well as medications. Our initial models were adjusted for sex and ethnicity. After further adjustment for SES, BMI, smoking status, alcohol use, physical activity, total cholesterol, triglycerides, HbA1c, systolic blood pressure, antihypertensive and glucose lowering medication, the association between change in autonomic nervous function and the development of arterial stiffness was attenuated. This suggests that part of the association can be explained by confounding by these covariates, that contribute both to the development of autonomic dysfunction and to arterial stiffness.

In principle, our models yield separate estimates for the effects of HRV level and change. However, we believe that the results can only be understood by considering observed combinations of the two determinants. E.g. if we examined the independent association between HRV levels and PWV (at a fixed rate of annual HRV change), it would seem that lower HRV levels were associated with lower PWV. We believe that this is probably due to the very strong positive correlation between HRV levels and their annual change in the study population, meaning that the strongest annual decline in HRV occurs in those with lowest HRV levels. Analysis of the effect of levels at a fixed rate of change would thus attempt to isolate the effect of the variation in levels not explained by the strong effect of annual change. To avoid these issues, our results are presented for (modelled) typical individuals within the observed ranges and combinations of HRV level and annual change.

As the association between autonomic dysfunction and arterial stiffness has previously been described in patients with diabetes, it would be particularly relevant to investigate if an individual’s glycaemic state has a modifying effect on the association between changes in autonomic nervous function and arterial stiffness. Although the current study is a longitudinal study and the outcome trajectories were assessed after the exposure trajectories, we cannot draw definitive causal conclusions, as other unmeasured or unknown common causes could still confound our findings. We can also not fully discount the theoretical possibility that arterial stiffness may have caused changes in autonomic nervous dysfunction (reverse causation). However, no obvious biologically plausible mechanisms point in this direction.

### 4.1 Strengths and limitations

Our study contains a relatively large study population with repeated measurement of exposures, outcomes and covariates. This is a methodological strength in our study due to the possibility to examine how the individual level and change in autonomic nervous function contributes to the development of arterial stiffness. The benefit of using linear mixed effect models is that this approach does not require having the same number of measurements or same time of recordings for each participant and thus makes optimal use of all available data (39). However, in the current study, the two exposures (HRV level and change) are correlated.

Of the 10,308 participants, 3,896 were excluded due to missing data on HRV, either because of death, non-response or withdrawal. Of the 6,412 with HRV measured, 1,511 did not have any PWV assessment or complete information on covariates. However, this group did not differ from the study population with regards to general characteristics. Participants had to survive until phase 9 to be included. This may introduce healthy survivor bias which may have led to some underestimation of the association between HRV and PWV. Presumably, those who died before phase 9 had a steeper decrease in autonomic nervous function. Also, they might have higher level and increase in PWV.

The HRV index SDNN and LF mainly reflects the effect of both the parasympathetic and sympathetic function, whereas RMSSD and HF power mainly reflect parasympathetic activity (23). Some of these measures may best reflect longer-term HR variability patterns and hence require ECG traces covering a full or even multiple days. In addition, none of the available HRV indices reflect sympathetic activity independently. The short-term reproducibility of aortic PWV was documented in another study using the Whitehall II dataset with PWV as outcome. The authors invited 125 participants after phase 9 to undergo the entire clinical examination a second time within 60 days of the original examination. The examinations showed good reproducibility i.e. the median (25^th^ percentile, 75^th^ percentile) intra-individual difference in PWV was 0.08 m/s (−0.68 ; 0.93) (27).

The Whitehall II study is a UK-based occupational cohort, reflecting the constitution of the civil service in 1985. Women and non-white ethnic groups are underrepresented, placing some limitations on the generalizability of our results to wider populations.

## 5 Conclusion

In conclusion, our study suggests that among middle-aged and older adults decreasing autonomic nervous function is associated with higher levels of arterial stiffness. Our findings extend our understanding of the mechanisms involved in the development of CVD risk, by quantifying the association between the age-related decrease in autonomic nervous function and arterial stiffness.

## Data Availability

All data produced in the present study are available upon reasonable request to the authors

## Abbreviations

ECG: Electrocardiogram
CAN: Cardiovascular autonomic neuropathy
CVD: Cardiovascular disease
HF power: High-frequency power
HRV: Heart rate variability
LF power: Low-frequency power
PWV: pulse wave velocity
SDNN: The standard deviation of normal-to-normal R-R intervals
rHR: Resting heart rate
RMSSD: The root mean square of the sum of the squares of differences between consecutive normal-to-normal R-R intervals

## 6 Acknowledgements

We thank all participating women and men in the Whitehall II Study, as well as all Whitehall II research scientists, study and data managers and clinical and administrative staff who make the study possible. JS acknowledges support by Steno Diabetes Center Aarhus, which is partially funded by an unrestricted donation from the Novo Nordisk Foundation.

## 7 Authors’ contributions

Study concept and design: JS, DRW, AH, MSC, LB, DV, CSH. Contributed the data: DRW, AGT. Planning the statistical analysis: JS, DRW, AH, LB, MSC, DV. Conducted the statistical analysis: JS. All authors contributed to, critically revised, and approved the final version of the manuscript. JS is the guarantor of this work and holds the final responsibility for the decision to submit for publication.

## 8 Funding

The UK Medical Research Council (K013351, R024227), British Heart Foundation and the US National Institutes of Health (R01HL36310, R01AG013196) have supported collection of data in the Whitehall II study.

## 9 Ethics

The study was approval The UK NHS Health Research Authority London-Harrow ethics committee approved the study which was conducted in accordance with the Helsinki Declaration with written informed consent from all participants.

## 10 Conflicts of interests

All the authors declare that there is no duality of interest associated with their contribution to this manuscript.

## Availability of data and materials

Whitehall II data, protocols, and other metadata are available to bona fide researchers for research purposes. Please refer to the Whitehall II data sharing policy at https://www.ucl.ac.uk/epidemiology-health-care/research/epidemiology-and-public-health/research/whitehall-ii/data-sharing

## 8 Supplementary material

**Table 1S:** Person-specific levels and change of rHR and SDNN by age groups.

| **Table 1S: Person-specific levels and change of rHR and SDNN by age groups** |  |  |  |  |
| --- | --- | --- | --- | --- |
| <u>Exposure</u> | N | Npe | Levels | Annual change |
| rHR | 4901 | 10249 |  |  |
| Age > 65 | 2625 | - | 66.63 bpm (61.9; 72.1) | -0.10 (-0.12; -0.08)<sup>1</sup> |
| Age 65 - 70 | 1057 | - | 66.21 bpm (61.3; 71.6) | -0.10 (-0.13; -0.07)<sup>1</sup> |
| Age ≥ 70 | 1219 | - | 66.33 bpm (61.2; 72.1) | -0.10 (-0.13; -0.06)<sup>1</sup> |
| SDNN | 4901 | 10249 |  |  |
| Age > 65 | 2625 | - | 32.9 ms (28.5; 37.5) | -1.5 (-1.8; -1.2)<sup>2</sup> |
| Age 65 - 70 | 1057 | - | 30.0 ms (25.1; 35.2) | -1.5 (-1.9; -1.0)<sup>2</sup> |
| Age ≥ 70 | 1219 | - | 27.4 ms (22.4; 33.6) | -1.5 (-1.9; -1.0)<sup>2</sup> |
| cSDNN | 4901 | 10249 |  |  |
| Age > 65 | 2625 | - | 3.6 (3.2; 4.1) | -1.6 (-1.9; -1.3)<sup>2</sup> |
| Age 65 - 70 | 1057 | - | 3.3 (2.8; 3.8) | -1.6 (-2.0; -1.2)<sup>2</sup> |
| Age ≥ 70 | 1219 | - | 3.0 (2.5; 3.6) | -1.6 (-2.0; -1.1)<sup>2</sup> |
| LF | 4901 | 10249 |  |  |
| Age > 65 | 2625 | - | 274.8 ms<sup>2</sup> (199.0; 381.6) | -3.7 (-4.4; -3.1)<sup>2</sup> |
| Age 65 - 70 | 1057 | - | 212.3 ms<sup>2</sup> (139.7; 309.4) | -3.7 (-4.5; -3.0)<sup>2</sup> |
| Age ≥ 70 | 1219 | - | 174.3 ms<sup>2</sup> (111.4; 276.3) | -3.7 (-5.0; -2.8)<sup>2</sup> |
| cLF | 4901 | 10249 |  |  |
| Age > 65 | 2625 | - | 3.3 (2.5; 4.6) | -4.0 (-4.7; -3.2)<sup>2</sup> |
| Age 65 - 70 | 1057 | - | 3.3 (2.8; 3.8) | -4.0 (-4.9; -3.2)<sup>2</sup> |
| Age ≥ 70 | 1219 | - | 3.0 (2.5; 3.6) | -3.9 (-4.8; -3.0)<sup>2</sup> |
| RMSSD | 4901 | 10249 |  |  |
| Age > 65 | 2625 | - | 20.0 ms (16.4; 24.0) | -1.2 (-1.7; -0.7)<sup>2</sup> |
| Age 65 - 70 | 1057 | - | 18.1 ms (14.4; 22.3) | -1.2 (-1.8; -0.7)<sup>2</sup> |
| Age ≥ 70 | 1219 | - | 17.1 ms (12.9; 23.4) | -1.2 (-1.8; -0.5)<sup>2</sup> |
| cRMSSD | 4901 | 10249 |  |  |
| Age > 65 | 2625 | - | 2.2 (1.9; 2.6) | -1.4 (-1.9; -0.9)<sup>2</sup> |
| Age 65 - 70 | 1057 | - | 2.0 (1.6; 2.4) | -1.4 (-1.9; -0.8)<sup>2</sup> |
| Age ≥ 70 | 1219 | - | 1.9 (1.5; 2.5) | -1.3 (-2.0; -0.6)<sup>2</sup> |
| HF | 4901 | 10249 |  |  |
| Age > 65 | 2625 | - | 110.6 ms<sup>2</sup> (76.2; 161.1) | -3.5 (-4.1; -2.9)<sup>2</sup> |
| Age 65 - 70 | 1057 | - | 87.5 ms<sup>2</sup> (56.3; 133.6) | -3.5 (-4.3; -2.7)<sup>2</sup> |
| Age ≥ 70 | 1219 | - | 72.0 ms<sup>2</sup> (43.4; 123.8) | -3.5 (-4.4; -2.4)<sup>2</sup> |
| cHF | 4901 | 10249 |  |  |
| Age > 65 | 2625 | - | 1.3 (0.9; 1.9) | -3.8 (-4.4; -3.0)<sup>2</sup> |
| Age 65 - 70 | 1057 | - | 1.0 (0.7; 1.6) | -3.8 (-4.3; -3.0)<sup>2</sup> |
| Age ≥ 70 | 1219 | - | 0.9 (0.5; 1.4) | -3.6 (-4.5; -2.8)<sup>2</sup> |
| Estimates represent each participant's rHR/SDNN level and annual change at age in phase 9<br>Results of the median with (25<sup>th</sup> percentile, 75<sup>th</sup> percentile)<br>N, number of participants used in particular analysis, Npe, number of person examinations used in particular analysis<br><sup>1</sup> Change in bpm per year<br><sup>2</sup> Percentage change in HRV per year |  |  |  |  |

**Table 2S:** Modelled 5-year trajectories of HRV or rHR association with annual PWV levels and change.

|  | Level of PWV<sup>1</sup> (m/s) |  | Annual change in PWV (m/s per year) |  |
| --- | --- | --- | --- | --- |
|  | Model 1<br>(n=4895) | Model 2<br>(n=4895) | Model 1<br>(n=4895) | Model 2<br>(n=4895) |
| **PWV by rHR** |  |  |  |  |
| **Age 60-65 with rHR level = 65** |  |  |  |  |
| 0.0 bpm rHR change per year | 7.81 (7.34; 8.28) | 7.72 (7.26; 8.18) | 0.12 (0.08; 0.16) | 0.13 (0.09; 0.17) |
| -0.2 bpm rHR change per year | 6.47 (6.03; 6.90) | 7.02 (6.60; 7.44) | 0.18 (0.15; 0.22) | 0.14 (0.11; 0.17) |
| Difference<sup>2</sup> | 1.34 (0.56; 2.13) | 0.70 (-0.03; 1.43) | -0.06 (-0.13; 0.2) | -0.01 (-0.07; 0.06) |
| **Age 65-70 with rHR level = 67** |  |  |  |  |
| 0.0 bpm rHR change per year | 8.57 (8.21; 8.93) | 8.47 (8.12; 8.83) | 0.13 (0.09; 0.16) | 0.13 (0.10; 0.17) |
| -0.2 bpm rHR change per year | 7.52 (7.15; 7.89) | 7.82 (7.45; 8.18) | 0.18 (0.14; 0.22) | 0.14 (0.10; 0.17) |
| Difference<sup>2</sup> | 1.05 (0.46; 1.63) | 0.65 (0.11; 1.20) | -0.06 (-0.13; 0.2) | -0.01 (-0.07; 0.06) |
| **Age 70-75 with rHR level = 70** |  |  |  |  |
| 0.0 bpm rHR change per year | 9.36 (9.05; 9.70) | 9.25 (8.92; 9.57) | 0.13 (0.10; 0.16) | 0.13 (0.10; 0.16) |
| -0.2 bpm rHR change per year | 8.61 (8.18; 9.04) | 8.64 (8.22; 9.05) | 0.19 (0.14; 0.23) | 0.14 (0.10; 0.18) |
| Difference<sup>2</sup> | 0.76 (0.16; 1.37) | 0.60 (0.05; 1.67) | -0.06 (-0.13; 0.2) | -0.01 (-0.07; 0.06) |
| **PWV by SDNN** |  |  |  |  |
| **Age 60-65 with SDNN level = 33** |  |  |  |  |
| -2% annual change in SDNN | 8.05 (7.72; 8.38) | 7.96 (7.64; 8.29) | 0.11 (0.09; 0.14) | 0.10 (0.07; 0.12) |
| -1% annual change in SDNN | 6.27 (5.92; 6.64) | 7.71 (7.37; 8.06) | 0.20 (0.18; 0.22) | 0.16 (0.15; 0.18) |
| Difference<sup>2</sup> | 1.78 (1.28; 2.28) | 1.04 (0.57; 1.50) | - 0.9 (-0.13; -0.05) | -0.07 (-0.10; -0.03) |
| **Age 65-70 with SDNN level = 30** |  |  |  |  |
| -2% annual change in SDNN | 8.48 (8.22; 8.74) | 8.39 (8.13; 8.66) | 0.13 (0.11; 0.15) | 0.11 (0.09; 0.13) |
| -1% annual change in SDNN | 7.16 (6.82; 7.50) | 7.71 (7.37; 8.06) | 0.22 (0.19; 0.24) | 0.18 (0.16; 0.20) |
| Difference | 1.32 (0.95; 1.69) | 0.68 (0.34; 1.02) | - 0.9 (-0.13; -0.05) | -0.07 (-0.10; -0.03) |
| **Age 70-75 with SDNN level = 27** |  |  |  |  |
| -2% annual change in SDNN | 9.04 (8.79; 9.29) | 8.95 (8.69; 9.21) | 0.14 (0.13; 0.16) | 0.13 (0.11; 0.14) |
| -1% annual change in SDNN | 8.16 (7.82; 8.50) | 8.61 (8.27; 8.95) | 0.23 (0.20; 0.26) | 0.19 (0.16; 0.23) |
| Difference<sup>2</sup> | 0.88 (0.55; 1.21) | 0.34 (0.03; 0.65) | - 0.9 (-0.13; -0.05) | -0.07 (-0.10; -0.03) |
| Fixed effect results of level and annual change with 95% confidence intervals of 5-year trajectories of modelled individuals at age 60, 65 and 70.<br>Model 1: Sex= Men, Ethnicity= White<br>Model 2: Sex=Male, Ethnicity=. White, SES= Professional/executive, BMI= 25, Smoking status= Non-smoker, Alcohol use= 8 units per week, Physical activity= 13 hours weekly of moderate to vigorous, Total cholesterol= 5.2mmol/L, Triglycerides= 1mmol/L, HbA1c= 5.6%, Systolic blood pressure= 124mmHg, Anti-hypertensive medication= Not using, Glucose lowering medication= Not using |  |  |  |  |
| <sup>1</sup>Level of PWV represent the participant modelled age at phase 9<br><sup>2</sup>PWV level and change difference between two groups with different exposure of change in rHR or SDNN |  |  |  |  |

**Figure 1S:**
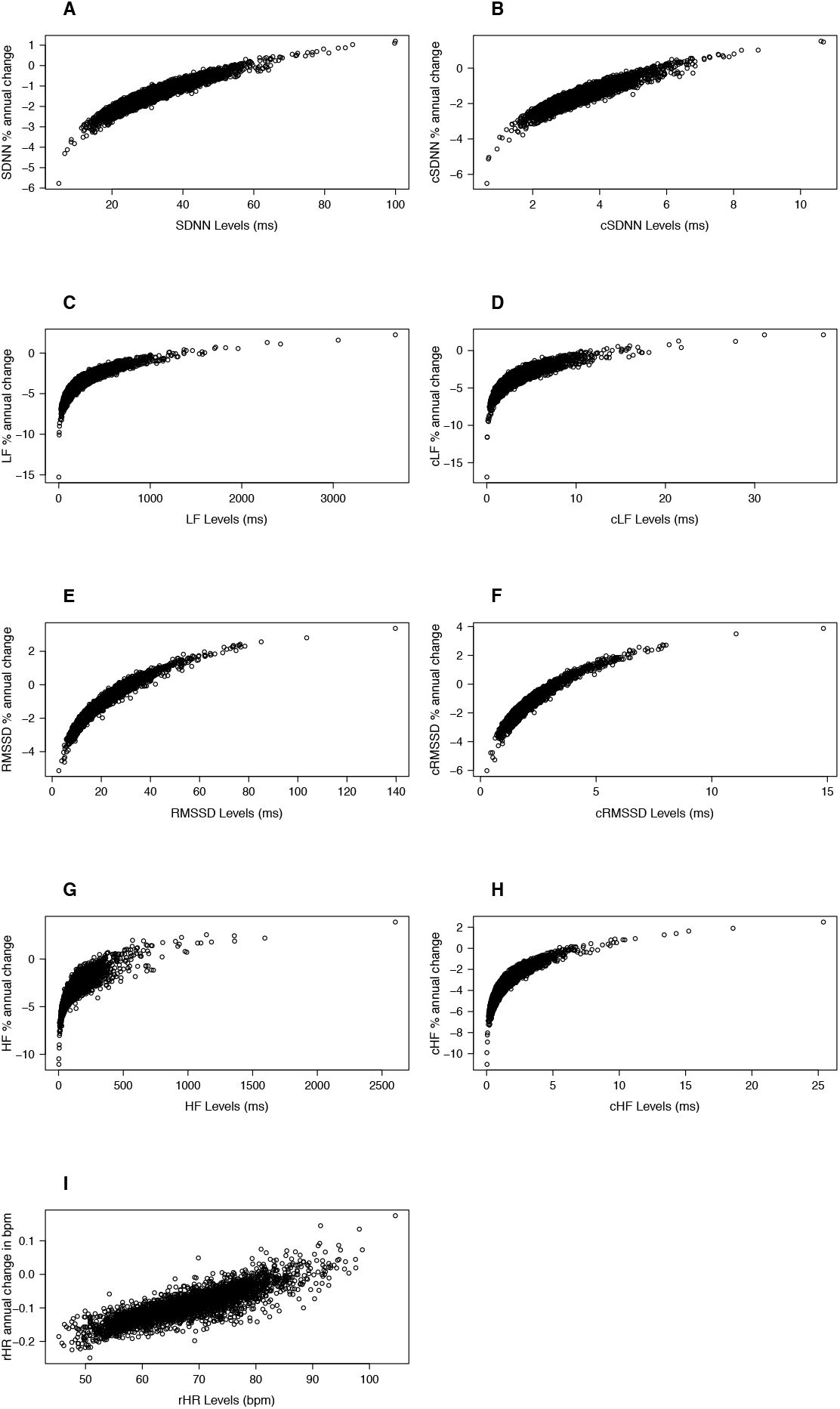
Plots of the study populations distribution of HRV/rHR level in phase 9 and the HRV percentage annual change/ rHR annual change in bpm. A) SDNN B) cSDNN C) LF D) cLF E) RMSSD F) cRMSSD G) HF H) cHF I) rHR

**Figure 2S:**
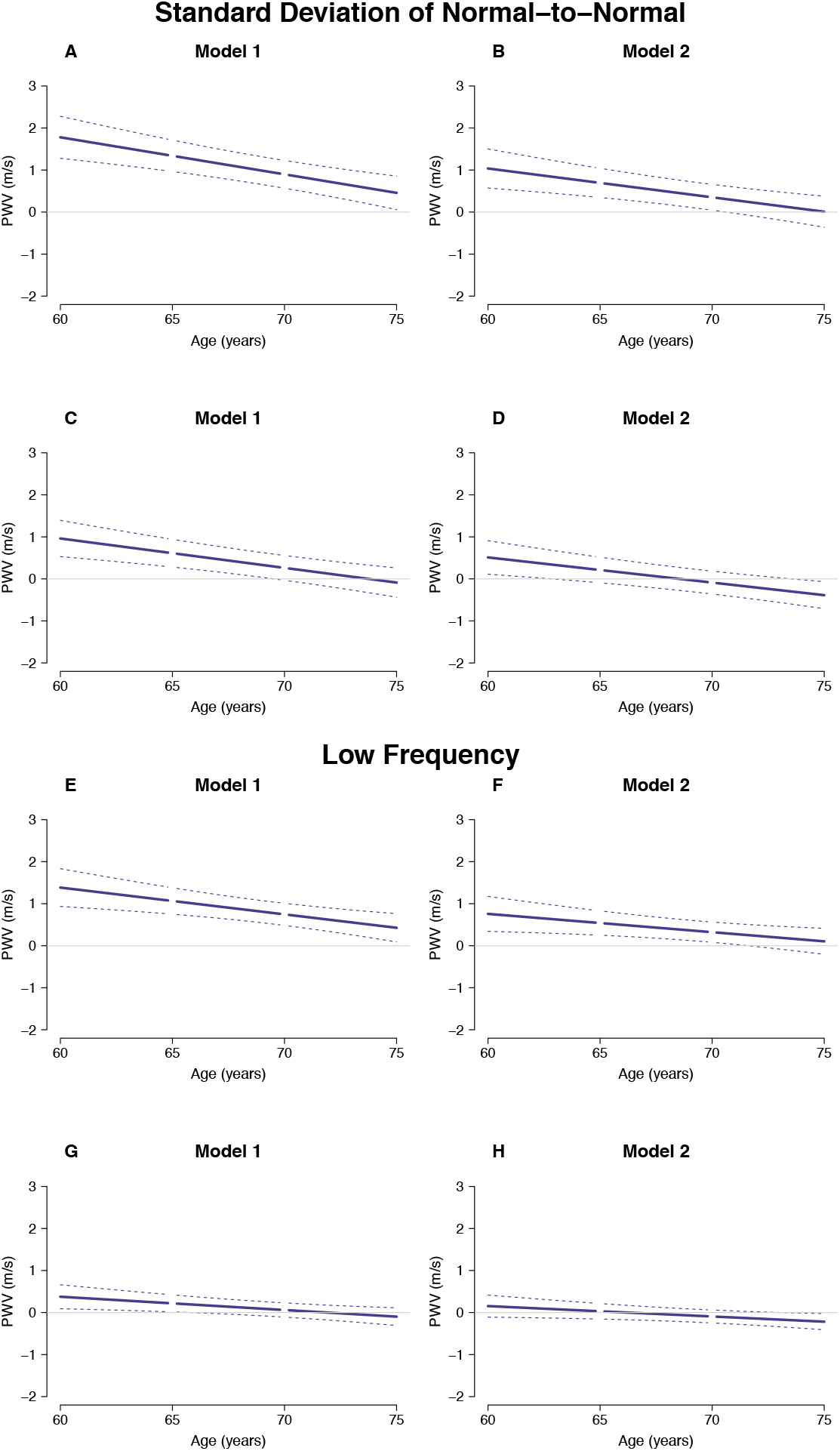
PWV difference by change in HRV indices that mainly are characterized by mixed sympathetic and parasympathetic influences (SDNN and LF) (difference in each panel in figure 2 in the article). A) Model 1: PWV difference between individuals with -1% and -2% annual SDNN decrease B) Model 2: PWV difference between individuals with -1% and -2% annual SDNN decrease C) Model 1: PWV difference between individuals with -1% and - 2% annual cSDNN decrease D) Model 2: PWV difference between individuals with -1% and -2% annual cSDNN decrease E) Model 1: PWV difference between individuals with -2% and -5% annual LF decrease F) Model 2: PWV difference between individuals with -2% and -5% annual LF decrease G) Model 1: PWV difference between individuals with -2% and -5% annual cLF decrease H) Model 2: PWV difference between individuals with -2% and -5% annual cLF decrease. The dotted lines show 95% confidence interval. Model 1 (Sex= Men, Ethnicity= White), Model 2 (Sex=Male, Ethnicity=. White, SES= Professional/executive, BMI= 25, Smoking status= Non-smoker, Alcohol use= 8 units per week, Physical activity= 13 hours weekly of moderate to vigorous, Total cholesterol= 5.2mmol/L, Triglycerides= 1mmol/L, HbA1c= 5.6%, Systolic blood pressure= 124mmHg, Antihypertensive medication= Not using, Glucose lowering medication= Not using)

**Figure 3S:**
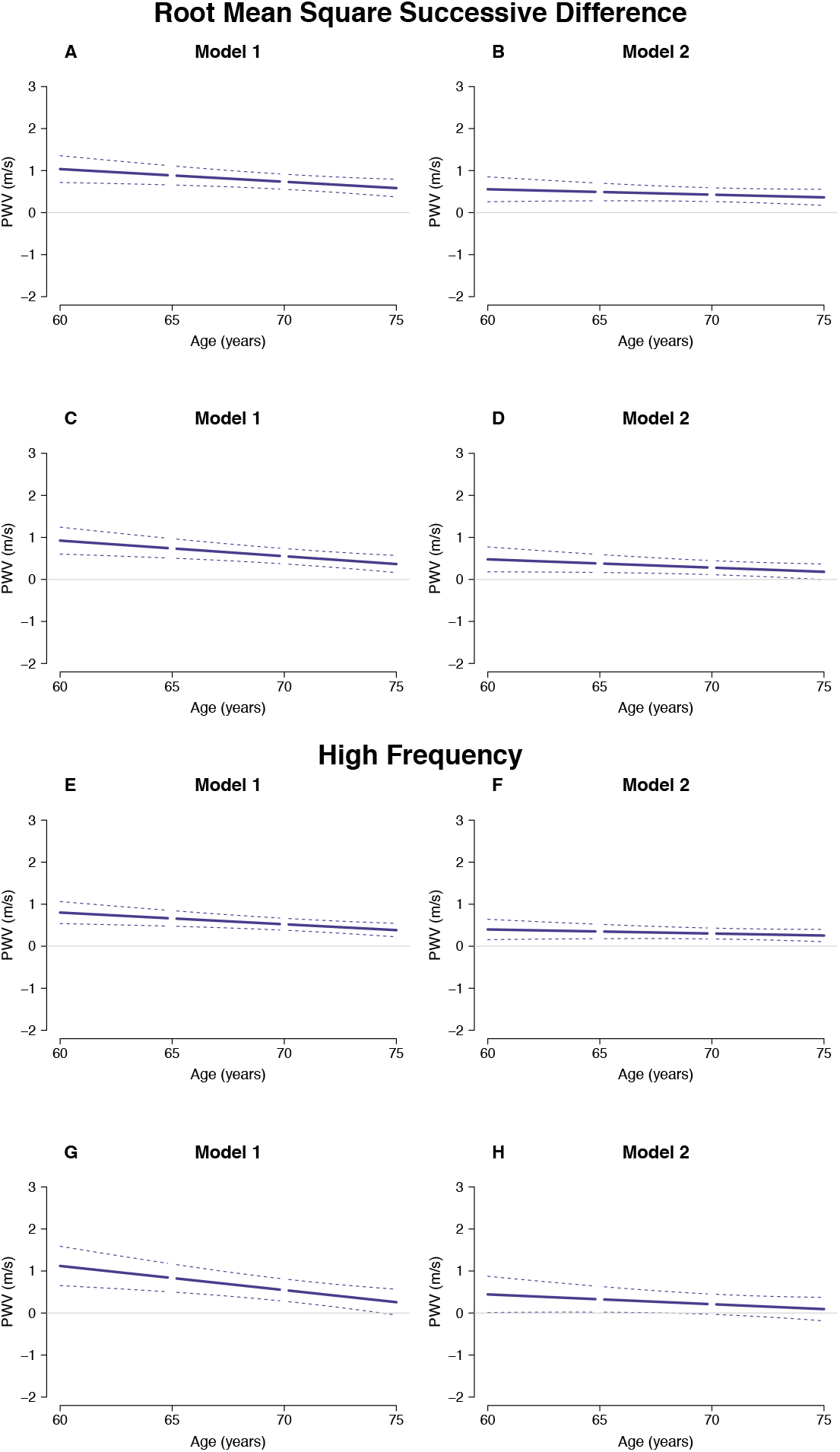
PWV difference by change in HRV indices that mainly are characterized by parasympathetic influence (RMSSD and HF) (difference in each panel in figure 3 in the article). A) Model 1: PWV difference between individuals with -1% and -2% annual RMSSD decrease B) Model 2: PWV difference between individuals with -1% and -2% annual RMSSD decrease C) Model 1: PWV difference between individuals with -1% and -2% annual cRMSSD decrease D) Model 2: PWV difference between individuals with -1% and -2% annual cRMSSD decrease E) Model 1: PWV difference between individuals with -2% and -4% annual HF decrease F) Model 2: PWV difference between individuals with -2% and -4% annual HF decrease G) Model 1: PWV difference between individuals with -2% and -5% annual cHF decrease H) Model 2: PWV difference between individuals with -2% and -5% annual cHF decrease. *The dotted lines show 95% confidence interval*. Model 1 (Sex= Men, Ethnicity= White), Model 2 (Sex=Male, Ethnicity=. White, SES= Professional/executive, BMI= 25, Smoking status= Non-smoker, Alcohol use= 8 units per week, Physical activity= 13 hours weekly of moderate to vigorous, Total cholesterol= 5.2mmol/L, Triglycerides= 1mmol/L, HbA1c= 5.6%, Systolic blood pressure= 124mmHg, Antihypertensive medication= Not using, Glucose lowering medication= Not using)

**Figure 4S:**
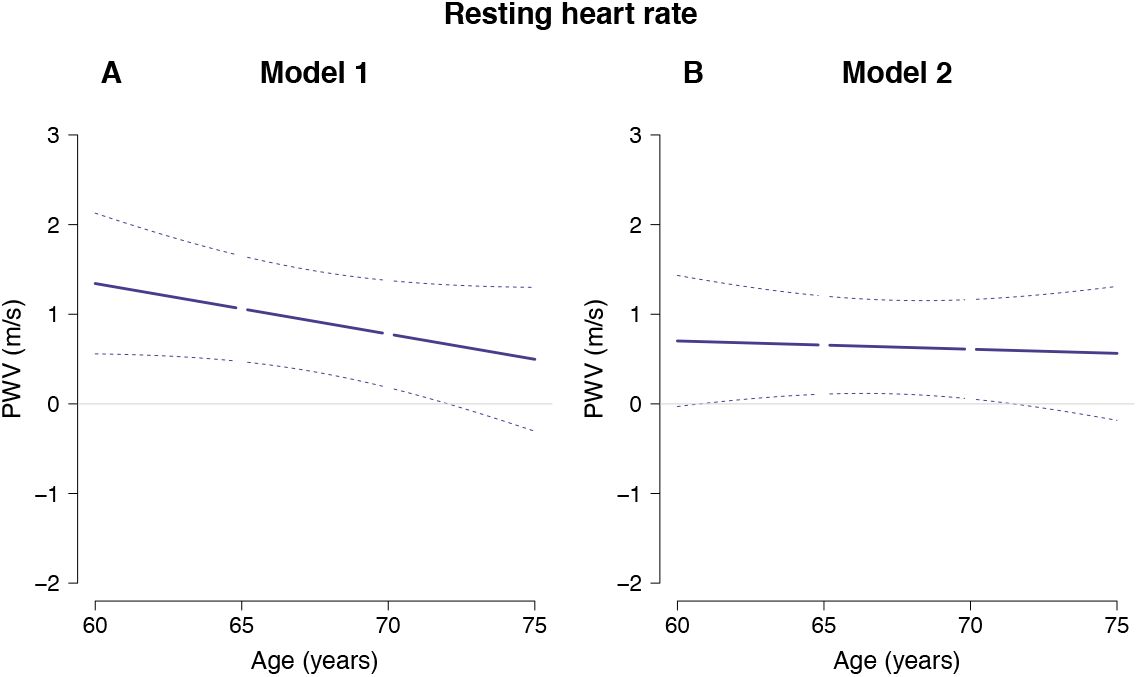
PWV difference by change in rHR (difference in each panel in figure 4 in the article). A) Model 1: PWV difference between individuals with 0.0% and –0.2% annual rHR change B) Model 2: PWV difference between individuals with 0% and -0.2% annual rHR change. *The dotted lines show 95% confidence interval*. Model 1 (Sex= Men, Ethnicity= White), Model 2 (Sex=Male, Ethnicity=. White, SES= Professional/executive, BMI= 25, Smoking status= Non-smoker, Alcohol use= 8 units per week, Physical activity= 13 hours weekly of moderate to vigorous, Total cholesterol= 5.2mmol/L, Triglycerides= 1mmol/L, HbA1c= 5.6%, Systolic blood pressure= 124mmHg, Antihypertensive medication= Not using, Glucose lowering medication= Not using)

**Figure 5S:**
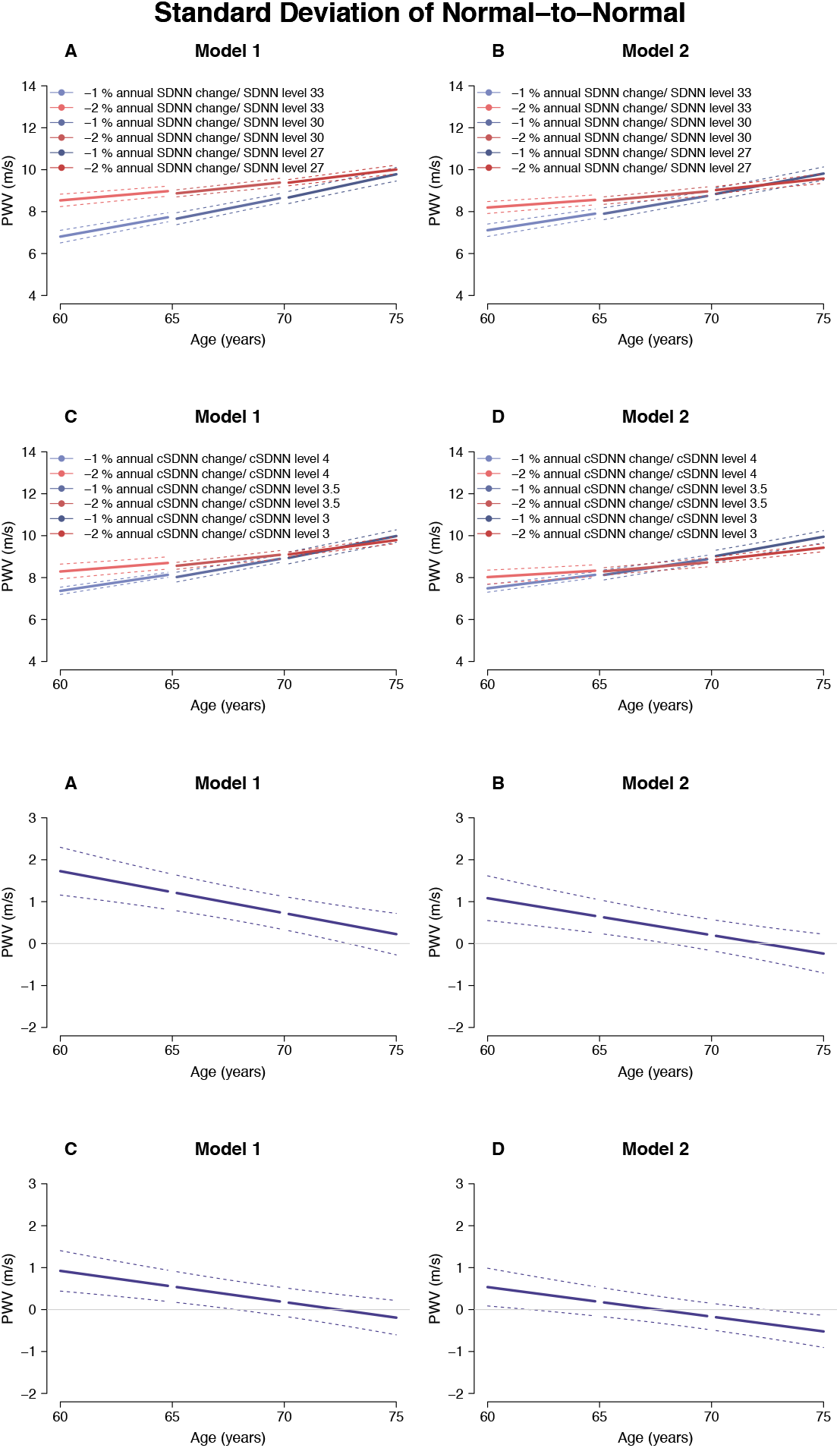
5-years PWV trajectories association with changes in SDNN in a subpopulation without diabetes. A) Model 1: PWV trajectories for individuals with either -1% or -2% annual SDNN decrease B) Model 2: PWV trajectories for individuals with either -1% or -2% annual SDNN decrease C) Model 1: PWV trajectories for individuals with either - 1% or -2% annual cSDNN decrease D) Model 2: PWV trajectories for individuals with either -1% or -2% annual cSDNN decrease. (The differences between the typical individuals are shown for the corresponding A, B, C, D figures below). *The dotted lines show 95% confidence interval*. Model 1 (Sex= Men, Ethnicity= White), Model 2 (Sex=Male, Ethnicity=. White, SES= Professional/executive, BMI= 25, Smoking status= Non-smoker, Alcohol use= 8 units per week, Physical activity= 13 hours weekly of moderate to vigorous, Total cholesterol= 5.2mmol/L, Triglycerides= 1mmol/L, HbA1c= 5.6%, Systolic blood pressure= 124mmHg, Antihypertensive medication= Not using, Glucose lowering medication= Not using)

### Estimation of heart rate adjusted HRV indices

Calculated interbeat interval (IBI) or heart period in milliseconds

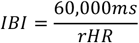

Adjusted SDNN indices

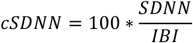

Adjusted RMSSD indices

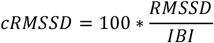

Adjusted HF indices

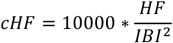

Adjusted LF indices

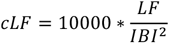

### Model specification by R-code using the nlme package

#### Step 1 analysis: Individual specific HRV levels and change

##### Non-adjusted HRV

lme (log(HRV) ∼ age, random = age | ID, method = “REML”, na.action = na.omit, data = Whitehall II)

##### Adjusted HRV

lme (log(cHRV) ∼ age, random = age | ID, method = “REML”, na.action = na.omit, data = Whitehall II)

##### Resting heart rate

lme (rHR ∼ age, random = age | ID, method = “REML”, na.action = na.omit, data = Whitehall II)

#### Step 2 analysis: Association between HRV level and change and the development of PWV

##### Non-adjusted HRV

*Model 1:* lme(PWV ∼ age * HRV intercept + age * HRV slope + sex + ethnicity, random = 1 | ID, method = “REML”, na.action = na.omit, data = Whitehall II)

*Model 2:* lme(PWV ∼ age * HRV intercept + age * HRV slope + sex + ethnicity + socio-economic status + BMI + smoking status + alcohol use + physical activity + total cholesterol + triglycerides + Hba1c + systolic blood pressure + anti-hypertensive medication + antidiabetic medication, random = 1 | ID, method = “REML”, na.action = na.omit, data = Whitehall II)

##### Adjusted HRV

*Model 1*: lme(PWV ∼ age * cHRV intercept + age * cHRV slope + sex + ethnicity, random = 1 | ID, method = “REML”, na.action = na.omit, data = Whitehall II)

Model 2: lme(PWV ∼ age * cHRV intercept + age * cHRV slope + sex + ethnicity + socio-economic status + BMI + smoking status + alcohol use + physical activity + total cholesterol + triglycerides + Hba1c + systolic blood pressure + anti-hypertensive medication + antidiabetic medication, random = 1 | ID, method = “REML”, na.action = na.omit, data = Whitehall II)

##### Resting heart rate

*Model 1*: lme(PWV ∼ age * rHR intercept + age * rHR slope + sex + ethnicity, random = 1 | ID, method = “REML” na.action = na.omit data = Whitehall II)

Model 2: lme(PWV ∼ age * rHR intercept + age * rHR slope + sex + ethnicity + socio-economic status + BMI + smoking status + alcohol use + physical activity + total cholesterol + triglycerides + Hba1c + systolic blood pressure + anti-hypertensive medication + antidiabetic medication, random = 1 | ID, method = “REML”, na.action = na.omit, data = Whitehall II)

## Notes

### Competing Interest Statement

The authors have declared no competing interest.

### Funding Statement

This study was funded by Steno Diabetes Center Aarhus, which is partially funded by an unrestricted donation from the Novo Nordisk Foundation

